# Factors associated with the clinical outcomes of maternal Intensive Care Unit admissions following obstetric surgeries at Muhimbili National Hospital, Tanzania: a cross-sectional study

**DOI:** 10.64898/2026.11.08.26367571

**Authors:** F.S. Ringo, M.M. Ulisubisya, R. Boniface, S. Mrutu, H. Msanga, A. Jongo, M.A. Zumba

## Abstract

Maternal ICU admission provides advanced, lifesaving medical care for pregnant or postpartum women who are critically ill. The burden of obstetric critical illness requiring ICU admission is higher in low- and middle-income countries (LMICs) than in high-income countries. Most critically ill obstetric patients admitted to the ICU have undergone major surgical interventions such as caesarean sections, hysterectomies and explorative laparotomies. Common indications for maternal ICU admission following surgery include postpartum hemorrhage (PPH), hypertensive disorders of pregnancy, and sepsis. Maternal ICU admission following obstetric surgery is associated with high maternal mortality, prolonged ICU stay, and poor neonatal outcomes. Identifying factors associated with poor maternal outcomes after ICU admission following obstetrics surgeries is essential for reducing preventable severe morbidity and mortality. This study aimed to assess the factors associated with the clinical outcomes of maternal ICU admissions following obstetric surgery at Muhimbili National Hospital (MNH), Dar es Salaam, Tanzania. This was a prospective cross-sectional study, conducted at the MNH, Obstetrics ICU, involving 115 women admitted following obstetric surgery. Data were collected using a modified WHO maternal near-miss checklist. Data were analyzed using SPSS version 27. Categorical variables were summarized using frequencies and percentages, and continuous variables were summarized using median and interquartile range (IQR). The associations between variables were assessed using Pearson’s chi-square and Fisher’s exact tests. Modified Poisson regression analysis was used to determine the strength of associations, with statistical significance set at p < 0.05 and 95% confidence interval. PPH was the most common indication for maternal ICU admission following surgery, accounting for 46.1% of admissions. The overall maternal ICU mortality was 16.5%, whereas 40.9% of patients experienced prolonged ICU stays. In bivariate analysis, mechanical ventilation (p = 0.04) and use of inotropes and/or vasopressors (p = 0.022) were significantly associated with maternal mortality, while mechanical ventilation (p = 0.025), PPH (p = 0.031), and blood and/or blood products transfusion (p = 0.003) were significantly associated with prolonged ICU stay. In the modified Poisson regression analysis, mechanical ventilation (adjusted prevalence ratio [aPR] = 2.3; 95% CI: 1.11–4.64; p = 0.025) and blood and/or blood product transfusion (aPR = 1.9; 95% CI: 1.23–2.80; p = 0.003) were independently and significantly associated with prolonged ICU stay.

This study demonstrates that PPH was the most common indication for maternal ICU admission following obstetric surgery. The overall maternal mortality was 16.5% with 40.9% of patients experience prolonged ICU stay. The need for mechanical ventilation, vasoactive support and blood transfusion were significantly associated with poor maternal outcomes. Healthcare providers should strengthen antenatal identification and correction of risk factors for PPH, including anaemia, previous PPH, multiple pregnancy, and hypertensive disorders of pregnancy during antenatal care visits (ANC); ensure appropriate prophylactic uterotonics administration during delivery; and promote early detection and timely, multidisciplinary management of PPH to reduce severe maternal morbidity, ICU admission, and maternal mortality. In addition, hospital management should strengthen early recognition and timely management of critically ill obstetric patients to reduce maternal mortality and prolonged ICU stay.

## INTRODUCTION

Maternal ICU admission provides advanced, lifesaving medical care for pregnant or postpartum women facing severe, life-threatening complications or who are critically ill (1). Maternal ICU admission is a critical sign used to measure maternal morbidity and is applicable in local surveillance and quality assurance activities related to pregnancy and puerperium (1) (2). The burden of obstetric critical illness requiring ICU admission is higher in LMICs than in high-income countries. The higher burden of critically ill obstetric patients requiring ICU admission in LMICs compared with HICs may be attributed to a higher burden of severe obstetric complications, delayed healthcare-seeking and referral, limited access to timely emergency obstetric care, inadequate critical-care infrastructure and resources, and limited availability of intermediate-care facilities (3). Most critically ill obstetric patients admitted to the ICU have undergone major surgical interventions such as cesarean sections, hysterectomies and explorative laparotomies (4).

Maternal ICU admission after obstetric surgery can occur due to multiple indications, which are pregnancy and non-pregnancy-related. According to the WHO, 10% of women who undergo CS will need exceptional care in a high-dependency unit (HDU) during the postoperative period, and ICU transfers account for 3.5% of them (5). Common reasons for maternal ICU admission following obstetric surgeries include, PPH, hypertensive disorders of pregnancy and sepsis (6). In Sub-Saharan Africa, PPH, eclampsia, sepsis and septic shock are the most common indications for ICU admission following obstetric surgeries (7–9). General anesthesia-related complications, such as difficult or failed intubation and anaphylaxis during obstetric surgery, may result in maternal ICU admission (10). High spinal and total spinal anaesthesia are other anesthesia-related complications that may occur during obstetric surgery and may cause maternal ICU admission (11).

Following maternal ICU admission after obstetric surgeries, different outcomes may occur for both the mother and the baby. Globally, maternal mortality among women admitted to ICU following obstetric surgeries varies substantially across settings, with considerably higher mortality reported in LMICs than in HICs. A recent systematic review reported pooled ICU mortality of 12.4% in LMICs compared with 1.4% in HICs (12–14). Maternal mortality following obstetric surgeries and ICU admission in Sub-Saharan Africa remains an urgent public health crisis. Data indicates that maternal mortality rates among obstetric patients admitted to ICUs after different surgical interventions ranges from 26% to 49% (15)(16). Prolonged ICU stay and adverse neonatal outcomes are among the reported outcomes associated with maternal ICU admission following obstetric surgery (17)(18).

In Tanzania, different studies have shown multiple factors associated with Maternal ICU admission after obstetric surgeries and the complications encountered. According to a study conducted at Tumbi Regional Referral Hospital, PPH was the most common indication for maternal ICU admission after cesarean section (19). From the study done at KCMC, preoperative anemia, grand multiparity, and longer surgical times were linked to more complications, which result in maternal ICU admission post-cesarean section (20).

Although MNH has a separate maternal ICU to treat critically ill obstetric patients admitted from the hospital’s maternal wards, theatres, and referrals from other hospitals, little is known about the factors that influence maternal ICU admission and the clinical outcomes following obstetric surgeries.

## METHODS

### Study design and settings

This study employed a prospective cross-sectional study design that aimed at assessing factors associated with the clinical outcomes of maternal intensive care unit admission following obstetric surgeries at MNH, Tanzania. This study was conducted in the Obstetric ICU of MNH-Upanga in Dar es Salaam, Tanzania. MNH serves as a National Referral Hospital, Research Center, and University teaching Hospital with a 1,500-bed capacity and 2,000 outpatients per day. MNH provides comprehensive maternal healthcare services and has a dedicated ICU for obstetric patients, making it an ideal site for evaluating maternal ICU admissions following obstetric surgeries. The maternal ICU has eight beds, each with a bedside monitor and a working mechanical ventilator. Twenty-three Nurses are stationed in the unit with different levels of education (Master’s degree -1, Degree 6, Diploma -10, Attendants-6). There are two stationed registrars, three intensivists and obstetricians on duty who normally perform two ward rounds per day (morning and evening rounds).

### Study period

The study was conducted over a six-month period, from 08/05/2025 to 08/11/2025.

### Study population

The study involved all women who were admitted to the Obstetric ICU at MNH following obstetric surgeries from May to November 2025.

### Sample size and sampling technique

The sample size was estimated from Taro Yamane Formula of 1967

(21), n = N/1+N (E) ²

Whereby;

n = sample size

N = finite population

E = level of significance

1 = a constant value

From the MNH maternal ICU, on average, 15 patients were admitted following obstetric surgery (from the pilot study, which reviewed maternal ICU admissions from July 2024 to January 2025). Using this number to approximate admissions per six months, the finite population will be 105.

Therefore,

N = 105

E = 0.05

n = 105/1 + 105(0.05)²

n = **83.**

The sample size estimated using the Taro Yamane formula was 83; Since the study used consecutive enrolment of all eligible admission over the study period, and prior data suggested atleast 105 such admission would occur, thus consecutive approach was expected to exceed the calculated minimum, which it did with **115** patients enrolled.

A consecutive sampling technique was employed to obtain the calculated sample size. This technique involved identifying eligible participants as they were admitted to the obstetric ICU following obstetric surgeries during the study period, ensuring that all available and eligible cases were included until the target sample size was achieved.

### Inclusion criteria

1. Women admitted to the maternal MNH ICU following obstetric surgeries from May to November 2025 (including referrals from other hospitals).
2. Complete medical records of the participant

### Exclusion criteria

1. Incomplete medical records

### Data collection methods, tool and procedures

Data were collected by the principal investigator and a trained research assistant. The research assistant was trained on the study topic, data collection tool (a modified WHO near-miss tool), and procedures for obtaining informed consent. The research assistant supported the principal investigator in data collection during periods when the principal investigator was not present in the maternal ICU at MNH.

Screening for eligible participants was conducted daily in the maternal ICU to identify patients who had either died or been discharged from the ICU following obstetric surgery. Eligible participants were screened using the inclusion criteria. Informed consent was obtained from patients who were discharged alive, and for patients who died, consent was obtained from the next of kin.

The WHO Near-Miss Tool, originally developed to assess severe maternal complications and near-miss cases, was modified according to the study objectives and setting. The modified tool was reviewed by senior anesthesiologists and intensivists to ensure its relevance, clarity, and appropriateness for data collection among maternal ICU patients following obstetric surgery.

Once eligibility was confirmed, demographic and clinical information were extracted simultaneously from the patient’s medical records, ICU registers, anaesthesia notes, surgical notes, and ICU management charts using the study data collection tool. collected information included sociodemographic, obstetric, and surgical characteristics; type of anaesthesia; category of surgery; indications for ICU admission; intraoperative complications; and interventions performed in the ICU, such as mechanical ventilation, blood transfusion, and use of inotropes and/or vasopressors. Outcome variables, including discharge status (alive or dead) and duration of ICU stay, were obtained from the same hospitalization records and documented accordingly.

### Data analysis

Data were entered and analyzed using the Statistical Package for the Social Sciences (SPSS) version 27. Descriptive statistics were used to summarize the participants’ demographic and clinical characteristics. Categorical variables were summarized using frequencies and percentages, and continuous variables were summarized using median and interquartile range (IQR). The indications for maternal ICU admission were summarized using frequencies and percentages. Pearson’s Chi-square test was used to assess the association between patient characteristics and outcomes (mortality and prolonged ICU stay). Fisher’s exact test was applied when the expected cell counts were less than five. Statistical significance was set at p < 0.05. Subsequently, modified Poisson regression was performed with univariate analysis to identify independent variables with p-values <0.25 as candidates for multivariate analysis to adjust for potential confounders. In the multivariate analysis, a p-value <0.05 was considered statistically significant, and the prevalence ratio was used to measure the strength of association.

### Ethical issues and research clearance

Ethical approval for the study was granted by the Muhimbili University of Health and Allied Sciences (MUHAS) Institutional Review Board (IRB) (Approval No. MUHAS-REC-04-2025-2788). Permission to conduct the study was also obtained from the Directorate of Research Services at Muhimbili National Hospital (MNH).

Written informed consent was obtained from patients who were discharged alive. For patients who had died, consent was sought from their next of kin, where feasible. Participants or their next of kin were provided with written information explaining the aim and significance of the study and the data collection procedures. Those who agreed to participate signed a written consent form. Participation was entirely voluntary, and participants were informed of their right to withdraw from the study at any time. Confidentiality was maintained throughout the study, and participants’ information was handled securely.

## RESULTS

### Social-demographics and clinical Characteristics

Table 1 present the Social-demographic and clinical characteristics of patient admitted to the Obstetric ICU. The majority of patients were aged 20–34 years (67.8%). Two-third of the patients (76.5%) were referred from other health facilities. Regarding antenatal care attendance, most patients (78.3%) attended four or more visits. In terms of parity, 61.7% of patients had low-risk parity, and 66.1% of patients delivered preterm. Most patients (67.0%) had no documented comorbidity. Regarding obstetric history, 33.0% of patients had a history of obstetric surgery. Additionally, complications during pregnancy were observed in 73.0% of patients.

**Table 1:**
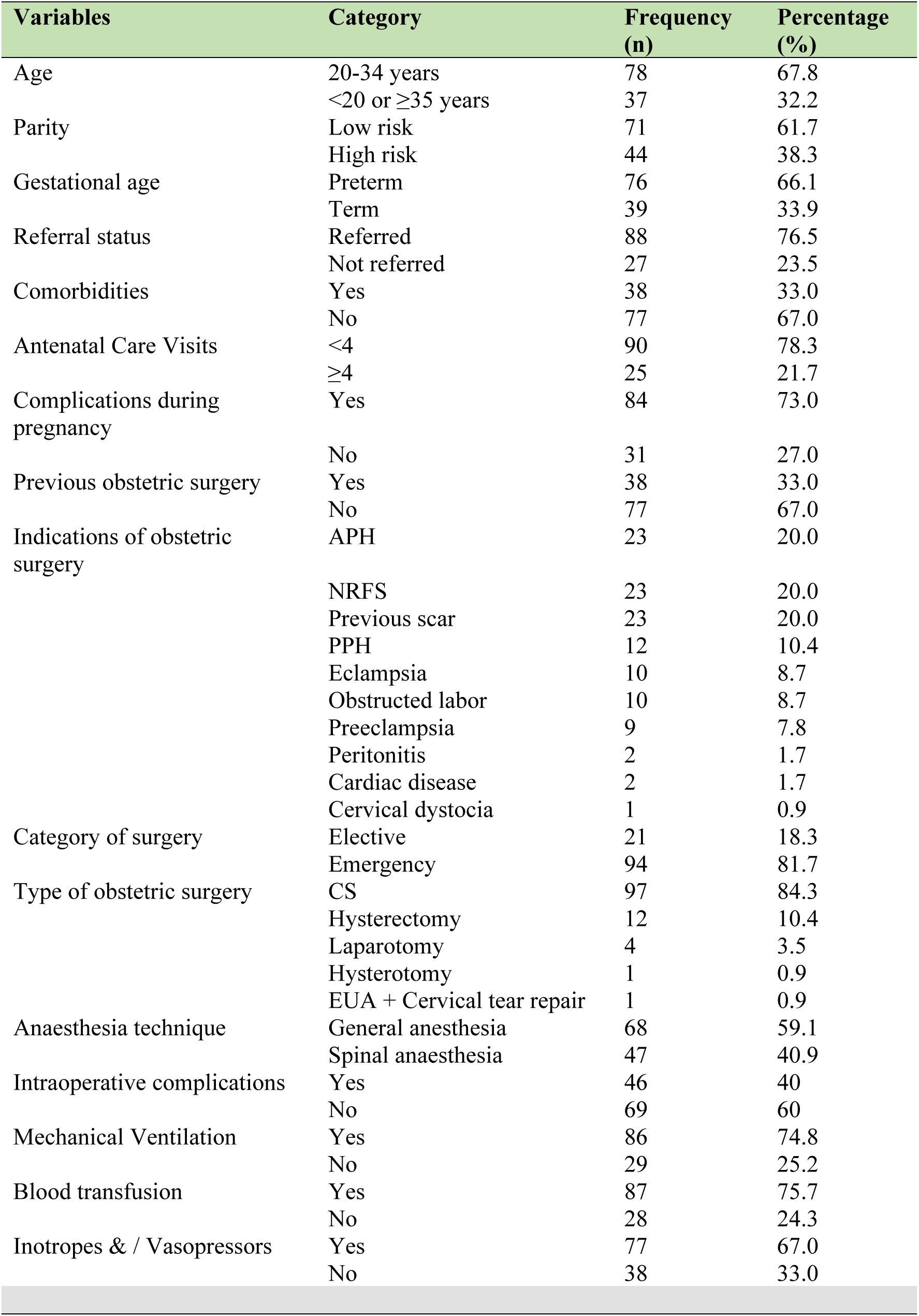
Sociodemographic and clinical characteristics of patients admitted to the maternal ICU at MNH following obstetric surgery, May–November 2025. (N=115)

| **Variables** | **Category** | **Frequency (n)** | **Percentage (%)** |
|---|---|---|---|
| Age | 20-34 years | 78 | 67.8 |
|  | <20 or ≥35 years | 37 | 32.2 |
| Parity | Low risk | 71 | 61.7 |
|  | High risk | 44 | 38.3 |
| Gestational age | Preterm | 76 | 66.1 |
|  | Term | 39 | 33.9 |
| Referral status | Referred | 88 | 76.5 |
|  | Not referred | 27 | 23.5 |
| Comorbidities | Yes | 38 | 33.0 |
|  | No | 77 | 67.0 |
| Antenatal Care Visits | <4 | 90 | 78.3 |
|  | ≥4 | 25 | 21.7 |
| Complications during pregnancy | Yes | 84 | 73.0 |
|  | No | 31 | 27.0 |
| Previous obstetric surgery | Yes | 38 | 33.0 |
|  | No | 77 | 67.0 |
| Indications of obstetric surgery | APH | 23 | 20.0 |
|  | NRFS | 23 | 20.0 |
|  | Previous scar | 23 | 20.0 |
|  | PPH | 12 | 10.4 |
|  | Eclampsia | 10 | 8.7 |
|  | Obstructed labor | 10 | 8.7 |
|  | Preeclampsia | 9 | 7.8 |
|  | Peritonitis | 2 | 1.7 |
|  | Cardiac disease | 2 | 1.7 |
|  | Cervical dystocia | 1 | 0.9 |
| Category of surgery | Elective | 21 | 18.3 |
|  | Emergency | 94 | 81.7 |
| Type of obstetric surgery | CS | 97 | 84.3 |
|  | Hysterectomy | 12 | 10.4 |
|  | Laparotomy | 4 | 3.5 |
|  | Hysterotomy | 1 | 0.9 |
|  | EUA + Cervical tear repair | 1 | 0.9 |
| Anaesthesia technique | General anesthesia | 68 | 59.1 |
|  | Spinal anaesthesia | 47 | 40.9 |
| Intraoperative complications | Yes | 46 | 40 |
|  | No | 69 | 60 |
| Mechanical Ventilation | Yes | 86 | 74.8 |
|  | No | 29 | 25.2 |
| Blood transfusion | Yes | 87 | 75.7 |
|  | No | 28 | 24.3 |
| Inotropes & / Vasopressors | Yes | 77 | 67.0 |
|  | No | 38 | 33.0 |

The most common indications for obstetric surgery were APH (20.0%), NRFS (20.0%), and previous scars (20.0%). Caesarean section was the most frequently performed surgical procedure (84.3%), and the majority of surgeries were conducted as emergency procedures (81.7%). General anesthesia was administered in 59.1% of the cases. Intraoperative complications were documented in 40% of the patients. With regard to ICU management, 74.8% of the patients required mechanical ventilation, 67.0% received inotropes and/or vasopressors support, and 75.7% underwent blood and/or blood products transfusion.

### Indications for maternal ICU admission

Table 2 presents the indications for maternal ICU admission after obstetric surgeries. The leading indication for ICU admission was PPH (46.1%), followed by preeclampsia (16.5%), eclampsia (14.8%), and sepsis (10.4%). Other indications included anesthesia-related complications (general anesthesia-related complications, high and total spinal anaesthesia) (5.2%) and DKA (1.7%). Less common indications were HEELP syndrome (0.9%), pulmonary embolism (0.9%), and cardiac disease (0.9%).

**Table 2:** Indications for maternal ICU admission at MNH following obstetric surgery, May-November 2025. (N=115)

| **Indication** | **Frequency (n)** | **Percentage (%)** |
|---|---|---|
| PPH | 53 | 46.1 |
| Preeclampsia | 19 | 16.5 |
| Eclampsia | 17 | 14.8 |
| Sepsis | 12 | 10.4 |
| Anaesthesia related complications | 9 | 7.8 |
| DKA | 2 | 1.7 |
| HEELP syndrome | 1 | 0.9 |
| Pulmonary Embolism | 1 | 0.9 |
| Cardiac disease | 1 | 0.9 |
| Total | N = 115 | 100% |

#### Clinical outcomes

Of the 115 patients included in the study, 96 (83.5%) were discharged alive, while 19 (16.5%) died during ICU admission, giving an overall maternal ICU mortality proportion of 16.5% (*Figure 1*). The median duration of ICU stay was 5 days (interquartile range, 3–8 days). Based on this median value, patients were categorized into short stay (≤5 days) and prolonged stay (>5 days), with 40.9% of patients experiencing a prolonged ICU stay *(Figure 2*).

**Figure 1:**
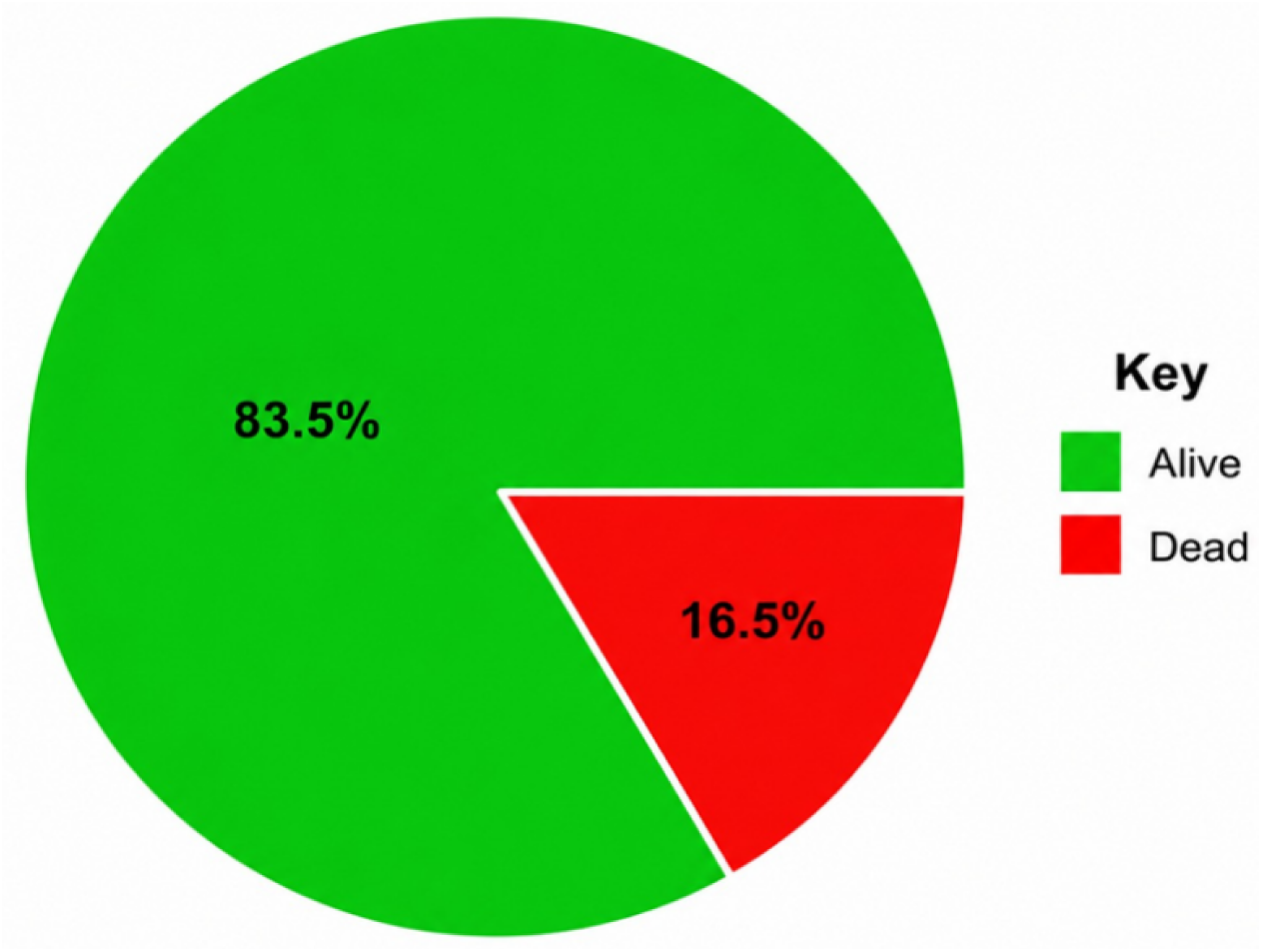
A Pie chart presentation of outcomes of patients admitted to the Obstetric ICU at MNH, following obstetric surgery, May - November 2025

**Figure 2:**
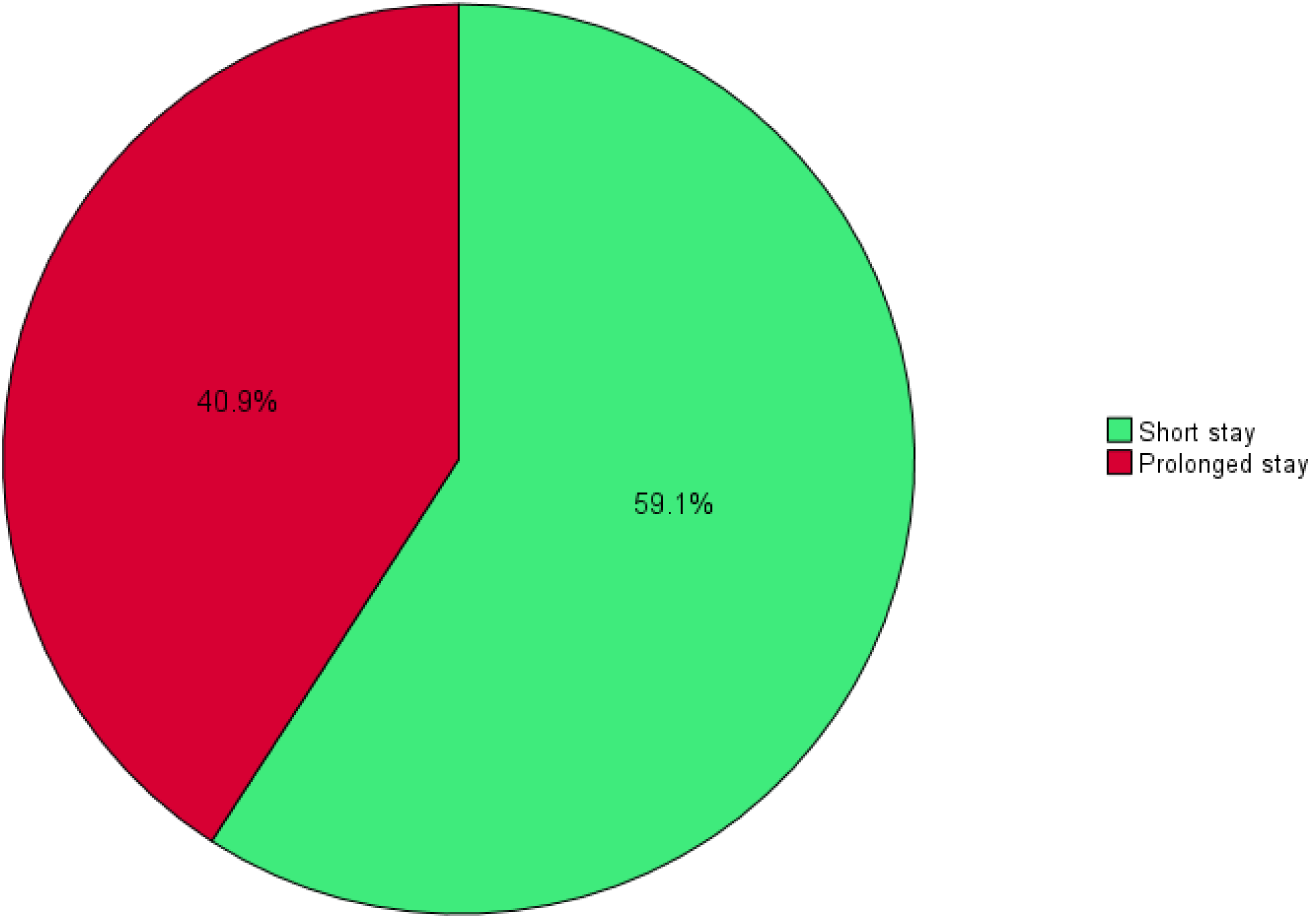
A pie chart presentation of duration of Obstetric ICU stay among patients admitted following obstetric surgery, May–November 2025

### Association between patient characteristics and clinical outcomes (Mortality and prolonged ICU stay)

#### Association between patient characteristics and mortality

Bivariate analysis was performed to assess these associations. Patients who required mechanical ventilation had significantly association with mortality than those who did not (p = 0.04). Similarly, the use of inotropes and/or vasopressors was significantly associated with mortality (p = 0.022). No statistically significant associations were observed between mortality and other variables, as shown in the table 3 below. On univariate and multivariate analysis (Modified Poisson regression), there were no factors which shows or retain their association with mortality.

**Table 3:** Association between patient characteristics and mortality among patients admitted to the maternal ICU at MNH following obstetric surgery, May - November 2025. (N=115)

| Variable | Category | Alive n (%) | Dead n (%) | p-value |
|---|---|---|---|---|
| Age group (years) | 20 - 34 | 65 (67.7) | 13 (68.4) | 0.952 |
|  | <20 or ≥35 | 31 (32.3) | 6 (31.6) |  |
| Parity | Low risk | 63 (65.6) | 8 (42.1) | 0.054 |
|  | High risk | 33 (34.4) | 11 (57.9) |  |
| GA | Preterm | 65 (67.7) | 11 (57.9) | 0.409 |
|  | Term | 31 (32.3) | 8 (42.1) |  |
| Referral status | Referred | 72 (75.0) | 16 (84.2) | 0.556* |
|  | Not referred | 24 (25.0) | 3 (15.8) |  |
| Comorbidities | Yes | 31 (32.3) | 7 (36.8) | 0.700 |
|  | No | 65 (67.7) | 12 (63.2) |  |
| ANC Visits | <4 | 74 (77.1) | 16 (84.2) | 0.761* |
|  | ≥4 | 22 (22.9) | 3 (15.8) |  |
| Complications during pregnancy | Yes | 71 (74.0) | 13 (68.4) | 0.619 |
|  | No | 25 (26.0) | 6 (31.6) |  |
| Previous obstetric surgery | Yes | 33 (34.4) | 5 (26.3) | 0.495 |
|  | No | 63 (65.6) | 14 (73.7) |  |
| Category of surgery | Elective | 20 (20.8) | 1 (5.3) | 0.190* |
|  | Emergency | 76 (79.2) | 18 (94.7) |  |
| Type of obstetric surgery | CS | 80 (83.3) | 17 (89.5) | 0.733* |
|  | Others | 16 (16.7) | 2 (10.5) |  |
| Anaesthesia technique | GA | 57 (59.4) | 11 (57.9) | 0.905 |
|  | Spinal | 39 (40.6) | 8 (42.1) |  |
| Intraoperative complications | Yes | 37 (38.5) | 9 (47.4) | 0.473 |
|  | No | 59 (61.5) | 10 (52.6) |  |
| PPH | Yes | 48 (50.0) | 5 (26.3) | 0.053 |
|  | No | 48 (50.0) | 14 (73.7) |  |
| Hypertensive disorders | Yes | 30 (31.2) | 9 (47.4) | 0.175 |
|  | No | 66 (68.8) | 10 (52.6) |  |
| Sepsis | Yes | 8 (8.3) | 4 (21.1) | 0.11* |
|  | No | 88 (91.7) | 15 (78.9) |  |
| Anaesthesia related complications | Yes | 8 (8.3) | 1 (5.3) | 1.0* |
|  | No | 88 (91.7) | 18 (94.7) |  |
| Mechanical Ventilation | Yes | 68 (70.8) | 18 (94.7) | **0.040**\* |
|  | No | 28 (29.2) | 1 (5.3) |  |
| Blood and /Blood transfusion | Yes | 73 (76.0) | 14 (73.7) | 0.778* |
|  | No | 23 (24.0) | 5 (26.3) |  |
| Inotropes and /Vasopressor use | Yes | 60 (62.5) | 17 (89.5) | **0.022** |
|  | No | 36 (37.5) | 2 (10.5) |  |

Key: Pearson Chi- square; p-value <0.05, \*Fisher’s exact

#### Association between patient characteristics and prolonged ICU stay

Table 4 shows bivariate analysis of factors associated with prolonged ICU stay. The following factors were significantly associated with prolonged ICU stay; Mechanical ventilation (p = 0.011), PPH (p = 0.031), blood and/or blood product transfusion (p = 0.014). No statistically significant associations were identified between prolonged ICU stay and the rest of the variables.

**Table 4:** Association between patient characteristics and prolonged ICU stay among patients admitted to the maternal ICU at MNH following obstetric surgery, May - November 2025. (N=115)

| Variable | Category | No n (%) | Yes n (%) | p-value |
|---|---|---|---|---|
| Age group (years) | 20-34 | 45 (66.2) | 33 (70.2) | 0.649 |
|  | <20 or ≥35 | 23 (33.8) | 14 (29.8) |  |
| Parity | Low risk | 46 (67.6) | 25 (53.2) | 0.117 |
|  | High risk | 22 (32.4) | 22 (46.8) |  |
| GA | Preterm | 45 (66.2) | 31 (66.0) | 0.981 |
|  | Term | 23 (33.8) | 16 (34.0) |  |
| Referral status | Referred | 51 (75.0) | 37 (78.7) | 0.643 |
|  | Not referred | 17 (25.0) | 10 (21.3) |  |
| Comorbidities | Yes | 19 (27.9) | 19 (40.4) | 0.162 |
|  | No | 49 (72.1) | 28 (59.6) |  |
| ANC Visits | <4 | 52 (76.5) | 38 (80.9) | 0.576 |
|  | ≥4 | 16 (23.5) | 9 (19.1) |  |
| Complications during pregnancy | Yes | 47 (69.1) | 37 (78.7) | 0.254 |
|  | No | 21 (30.9) | 10 (21.3) |  |
| Previous obstetric surgery | Yes | 23 (33.8) | 15 (31.9) | 0.831 |
|  | No | 45 (66.2) | 32 (68.1) |  |
| Category of surgery | Elective | 15 (22.1) | 6 (12.8) | 0.205 |
|  | Emergency | 53 (77.9) | 41 (87.2) |  |
| Type of surgery | CS | 61 (89.7) | 36 (76.6) | 0.057 |
|  | Others | 7 (10.3) | 11 (23.4) |  |
| Anaesthesia technique | GA | 37 (54.4) | 31 (66.0) | 0.216 |
|  | Spinal | 31 (45.6) | 16 (34.0) |  |
| Intraoperative complications | Yes | 23 (33.8) | 23 (48.9) | 0.104 |
|  | No | 45 (66.2) | 24 (51.1) |  |
| PPH | Yes | 37 (54.4) | 16 (34.0) | **0.031** |
|  | No | 31 (45.6) | 31 (66.0) |  |
| Hypertensive disorders | Yes | 19 (27.9) | 20 (42.6) | 0.104 |
|  | No | 49 (72.1) | 27 (57.4) |  |
| Sepsis | Yes | 6 (8.8) | 6 (12.8) | 0.545* |
|  | No | 62 (91.2) | 41 (87.2) |  |
| Anaesthesia related complications | Yes | 6 (8.8) | 3 (6.4) | 0.736* |
|  | No | 62 (91.2) | 44 (93.6) |  |
| Mechanical ventilation | Yes | 45 (66.2) | 41 (87.2) | **0.011** |
|  | No | 23 (33.8) | 6 (12.8) |  |
| Blood and blood products transfusion | Yes | 57 (83.8) | 30 (63.8) | **0.014** |
|  | No | 11 (16.2) | 17 (36.2) |  |
| Inotropes and or Vasopressor use | Yes | 41 (60.3) | 36 (76.6) | 0.068 |
|  | No | 27 (39.7) | 11 (23.4) |  |

Key: Pearson Chi- square; p-value <0.05, \*Fisher’s exact

### Univariate and Multivariate analysis (Modified Poisson regression) of factors associated with prolonged ICU stay

#### Univariate modified Poisson regression analysis

In the univariate analysis, as shown in Table 5 below, PPH, mechanical ventilation, and blood and/or blood product transfusion were found to be significantly associated with prolonged ICU stay (p < 0.25). Patients with PPH were 1.7 times more likely to have prolonged ICU stays than those without PPH (cPR = 1.7; 95% CI: 1.03–2.67; p = 0.039). Patients who required mechanical ventilation were 2.3 times more likely to have prolonged ICU stay compared to those who did not require ventilation (cPR = 2.3; 95% CI: 1.09–4.86; p = 0.028). Similarly, patients who received blood and/or blood product transfusions were 1.8 times more likely to have prolonged ICU stay than those who did not receive transfusions (cPR = 1.8; 95% CI: 1.16–2.67; p = 0.008).

**Table 5:** Univariate and Multivariate analysis (Modified Poisson Regression) of factors associated with prolonged ICU stay among patients admitted to the maternal ICU at MNH following obstetric surgery, May – November 2025. (N=115)

| **Variable** | **Prolonged ICU stay** |  | **Prevalence Ratio (95% CI)** |  |  |  |
|---|---|---|---|---|---|---|
|  | Yes | No | Crude (95% CI) | p-value | Adjusted (95% CI) | p-value |
| Mechanical ventilation |  |  |  |  |  |  |
| Yes | 41(87.2) | 45(66.2) | 2.3(1.093,4860) | 0.028 | 2.3(1.108,4.641) | **0.025** |
| No | 6(12.8) | 23(33.8) | 1<sup>a</sup> | - | - | - |
| Blood transfusion |  |  |  |  |  |  |
| Yes | 30(63.8) | 57(83.8) | 1.8(1.162,2.668) | 0.008 | 1.9(1.232,2.798) | **0.003** |
| No | 17(36.2) | 11(16.2) | 1<sup>a</sup> | - | - | - |
| PPH |  |  |  |  |  |  |
| Yes | 16(34.0) | 37(54.4) | 1.7(1.026,2.674) | 0.039 | 1.5(0.913,2.455) | 0.11 |
| No | 31(66.0) | 31(45.6) | 1<sup>a</sup> | - | - | - |

**Key: Univariate significance; p-value < 0.25: Multivariate significance; p-value < 0.05: 1<sup>a</sup>; Reference category**

#### Multivariate modified Poisson regression analysis

In the multivariate analysis, as shown in Table 5, after adjusting for confounders, mechanical ventilation and blood and/or blood product transfusion retained their statistically significant association with prolonged ICU stay. Patients who required mechanical ventilation were 2.3 times more likely to have prolonged ICU stay compared to those who did not require mechanical ventilation (aPR = 2.3; 95% CI: 1.11–4.64; p = 0.025). Similarly, patients who received blood and/or blood product transfusions were 1.9 times more likely to have prolonged ICU stay than those who did not receive transfusions (aPR = 1.9; 95% CI: 1.23–2.80; p = 0.003).

## DISCUSSION

### Indications for maternal ICU admission following obstetric surgery

#### Postpartum Hemorrhage (PPH)

In this study, PPH was the leading indication for maternal ICU admission, accounting for nearly half of all cases (46.1%). This reflects the critical nature of severe obstetric bleeding, which can rapidly lead to hypovolemic shock, coagulopathy, and multiorgan failure, requiring ICU admission for close management and monitoring. This finding aligns with the study conducted in India by Ashakiran et al. (22), who found similar results, with PPH as the leading cause of maternal ICU admission (44.05%). However, these results contrast with those of a study from Nigeria by Tobi et al. (7), where PPH contributed to 6.3% of maternal ICU admission. Studies conducted in tertiary referral hospitals may report higher proportions due to the concentration of severe cases, while variations in the availability of blood transfusion services, timely obstetric interventions, and ICU admission criteria also play significant roles. PPH remains a major contributor to maternal ICU admissions following obstetric surgery, indicating gaps in pre-operative screening during ANC visits, early recognition and timely management of the condition. Strengthening ANC visits screening and detection of women at risk of PPH and early referral for care, intra and post-operative vigilance, early use of uterotonics and tranexamic acid, and improving access to blood transfusion services are essential to reduce the progression to severe hemorrhage and ICU admission.

#### Preeclampsia

Preeclampsia was the second most common indication for maternal ICU admission in our study, contributing to 16.5% of all cases admitted. This finding aligns with the study conducted in Nigeria by Asudo et al.(23), which reported similar results, where preeclampsia contributed to 20.1% of cases admitted to the ICU. This finding is contrary to the findings of Keizer et al. (24) in the Netherlands, where preeclampsia contributed to 62.0% (higher proportion compared to our findings) of all maternal cases admitted to the ICU. The discrepancy in findings could be accounted for by differences in study design, as the study by Keizer was a 12 years retrospective study, which used a larger sample size than our study. Clinically, these findings highlight the need to strengthen the early identification and optimal perioperative management of hypertensive disorders during pregnancy. Ensuring effective antenatal screening, timely blood pressure control, appropriate use of magnesium sulfate for seizure prophylaxis, and close intraoperative and postoperative monitoring are critical in preventing the progression to severe complications requiring ICU care.

#### Eclampsia

Eclampsia accounted for 14.8% of ICU admissions in this study, highlighting the continued burden of poorly controlled hypertensive disorders during pregnancy. This proportion is higher than that reported in Iran (5.7%) by Farzi et al. (25) but considerably lower than the findings from Nigeria (54.3%) by Tobi et al. (7), suggesting that differences in antenatal care coverage, timeliness of intervention, and ICU admission practices may influence these variations. Clinically, these findings underscore the need to strengthen the early detection and effective management of hypertensive disorders before progression to eclampsia. Ensuring routine antenatal surveillance, timely initiation of antihypertensive therapy, and appropriate use of magnesium sulfate for seizure prevention are essential. Furthermore, improved perioperative monitoring, rapid stabilization of patients with severe preeclampsia, and timely surgical decision-making may reduce the progression to eclampsia and subsequent ICU admission.

#### Sepsis

Sepsis was another major indication for ICU admission in this study, accounting for 10.4% of all admitted patients, indicating its important contribution to postoperative maternal morbidity. This is somewhat consistent with a study conducted in the United Kingdom by Acosta et al. (26), where sepsis accounted for 14.4% of maternal ICU admissions. However, the findings of the current study differ from those of a study conducted in Portugal by Oliveira et al. (27), where a higher percentage (20.4%) of maternal ICU admissions due to sepsis was observed. Another study in South Africa by Lafon et al. (28) found that sepsis post-CS accounted for 50% of all patients admitted to the ICU. These discrepancies found in these studies could be attributed to the fact that Oliveira et al. involved both antepartum and postpartum patients admitted to the ICU, while Lafon et al. involved only patients with sepsis after CS, admitted to the ICU. Clinically, these findings emphasize the need for strict aseptic techniques, timely use of antibiotics, and close postoperative monitoring to prevent progression to severe sepsis, which requires ICU admission.

#### Anesthesia-related complications

Anesthesia-related complications were identified as a collective indication for ICU admission, encompassing complications from general anesthesia, total spinal anesthesia, and high spinal anesthesia, which contributed to 7.8% of maternal ICU admissions. This is contrary to the study conducted in Australia by Stephen et al. (29), where 12.6% of maternal ICU admissions were due to anesthesia-related complications. However, a study from Columbia by Carrasquilla et al. (30) reported a higher percentage (29.9%), similar to the study conducted in Nigeria by Amarengiaye et al. (31), where 50% of ICU admissions were due to general anesthesia-related complications. Discrepancies found in these studies could be attributed to the facts that, a study by Stephen et al was a 10 years retrospective study, with 126 cases involved, but Carrasquilla et al study involves only preeclampsia patient with severe criteria, however Amarengiaye et al study focused on anaesthesia related complications following CS necessitating ICU admission. Clinically, these findings highlight the importance of strengthening safe obstetric anaesthesia through adequate training, appropriate monitoring, and preference for safer techniques, such as regional anaesthesia, where feasible. Improving airway management skills, ensuring the availability of essential equipment, and enhancing perioperative vigilance are critical for reducing anesthesia-related complications and subsequent ICU admissions.

#### Diabetic Ketoacidosis (DKA)

Although relatively uncommon, diabetic ketoacidosis represents a serious metabolic indication for maternal ICU admission. It is frequently associated with precipitating factors such as infection and the physiological stress of delivery, including CS, and often requires intensive monitoring and critical-care support. In this study, DKA accounted for 1.7% of maternal ICU admissions. Similarly, a study conducted in California by Hersh et al. (32) found that 2.0% of patients with type 1 diabetes mellitus and DKA during pregnancy were admitted to the ICU. This study by Hersh et al. was an 11 years retrospective-cohort that involved 786 patients with type 1 diabetes mellitus and DKA, whereby DKA was reported to be one of the causes of maternal ICU admission during pregnancy. Clinically, these findings highlight the importance of optimal glycemic control, early identification of high-risk patients, and close perioperative monitoring of these patients. Prompt recognition and timely management of metabolic derangements are essential to prevent the progression to severe illness requiring ICU admission.

#### HELLP Syndrome

HELLP syndrome was a rare indication for ICU admission in this study, contributing to 0.9% of cases admitted to the maternal ICU. This finding aligns with the study conducted in Iran by Farzi et al. (25), where 4.1% of maternal ICU admissions were due to HELLP syndrome. Contrary to a study conducted in Turkey by Agacayak et al. (33), 9.4% of patients with HELLP syndrome were admitted to the ICU. Discrepancies in these studies could be attributed to the fact that the study by Farzi et al. involved both patients who were post-CS (90%) and those who were post-normal vaginal delivery, but a study by Agacayak et al. involved 83.7% of patients post-CS with HELLP syndrome, of which 32 patients were admitted to the ICU. Although relatively uncommon, HELLP syndrome is a life-threatening condition associated with significant maternal complications and ICU admissions. Clinically, these findings highlight the importance of early recognition of severe preeclampsia, timely intervention, and close perioperative monitoring to prevent the progression to HELLP syndrome and reduce ICU admissions.

#### Pulmonary embolism

Pulmonary embolism was an infrequent indication, contributed to 0.9% of maternal ICU admission following obstetric surgery. This finding aligns with the study conducted in Korea by Yi et al. (34), whereby 2.2% of maternal ICU admissions were due to pulmonary embolism. However, the findings of the current study differ from those of a study conducted in Austria by Farr et al. (35)in which 4.2% of maternal ICU admissions were due to pulmonary embolism, and from those of another study in Italy by Vargas et al. (36), in which pulmonary embolism accounted for 7% of maternal ICU admissions. These discrepancies could be attributed to the fact that, Yi et al involved 176-sample size, while Farr et al involved 238-sample size, which with 84.5% of cases underwent CS, but Vargas et al involved 66 cases of which 95% were post CS. Clinically, these findings emphasize the importance of routine risk stratification, use of thromboprophylaxis in high-risk patients, early mobilization, and prompt recognition and management to reduce ICU admissions and improve maternal outcomes

#### Cardiac disease

Cardiac disease was also a rare indication for maternal ICU admission in this study, contributing to 0.9% of all cases admitted to the maternal ICU. This proportion is lower than that reported by a study conducted in South Africa by Motiang et al. (37), in which 44% of ICU admissions were due to pre-existing cardiac disease. Similarly, a study conducted in North Carolina by Small et al. (38) reported that 36% of 86 women admitted to the ICU were due to cardiac disease. In addition, a study conducted in China by Zhang et al. (39) reported that 12% of maternal ICU admissions were due to cardiac disorders. Discrepancies observed in these studies could be attributed to the fact that Motiang et al. involved 210 women, delivered by CS (88.6%) and by normal vaginal delivery; Small et al. used a retrospective study design, while Zhang et al. involved 133 cases, of which 73.7% were post-CS. This wide variation likely reflects differences in the underlying prevalence of maternal heart disease, referral patterns, and ICU admission criteria across settings. Clinically, these findings highlight the importance of early identification of high-risk women with cardiac disease, multidisciplinary management, and careful perioperative monitoring to prevent decompensation and reduce the need for ICU admission.

### Clinical outcomes

#### Mortality

The maternal ICU mortality observed in this study was 16.5%, reflecting the significant burden of severe maternal complications among women admitted to the ICU after obstetric surgery. This finding is similar to other studies from LMICs; studies from Nigeria by Tobi et al. (7), demonstrated an overall ICU maternal mortality of 23.2%, from Malawi, by Prin et al. 41% (40) and from Ghana, by Anane-Fenin et al. 26% (41). However, studies from HICs, reported lower maternal mortality compared with these findings from LMICs. For instance, a study from Netherlands by Zwart el al. (42) reported maternal mortality of 3.5% and a study from United Kingdom by Harrison et al. (43)reported maternal mortality of 4.2%. The higher maternal mortality in LMICs compared with HICs is multifactorial, reflecting inadequate and delayed antenatal care and identification of high-risk pregnancies, delays in accessing and receiving skilled intrapartum and emergency obstetric care, and, following delivery or obstetric surgery, delayed ICU admission (for those who are critically ill) and limited availability of critical-care resources, including blood products, mechanical ventilation, vasoactive support and adequately trained multidisciplinary teams. Clinically, this mortality proportion underscores the need to strengthen the continuum of maternal care through improved ANC attendance and early identification of high-risk pregnancies, early recognition and timely management of obstetric complications, particularly at peripheral facilities, improving referral systems, ensuring rapid stabilization prior to transfer, and expanding access to critical care services are essential strategies for reducing maternal mortality.

#### Prolonged ICU stay

The median duration of ICU stay in this study was 5 days, with 40.9% of patients experiencing prolonged ICU stay (> 5 days), indicating a considerable burden on critical care resources. Evidence in the literature regarding duration of ICU stay post surgery is conflicting, a study from South Africa by Motiang et al. (37) reported that the median length of ICU stay was 24 hours. While a study conducted in Bangladesh by Khanum et al. (44)reported that most patients stayed in the ICU for up to two days. Discrepancies from these studies could be attributed to the fact that, in Motiang et al. study, most of the patients (87.1%) were hemodynamically stable and needed minimal intervention in the ICU, but Khanum et al. used a retrospective study design and only 105 patients were involved. The longer ICU stay observed in this study may be attributed to several factors, including delayed presentation, severity of obstetric complications such as hemorrhage and sepsis, and the need for prolonged organ support, such as mechanical ventilation and transfusion. However, a study from Turkey by Agacayak et al. (45)reported that the average length of stay in the ICU was 7 days due to transfusions requirements, which is higher than our study findings.

In this study, 40.9% of patient experienced prolonged ICU stay, similar to Wang et al. (46) reported that 39.5% of critically ill obstetric patients had a prolonged ICU stay. Clinically, prolonged ICU stay has important implications, including an increased risk of hospital-acquired complications, higher healthcare costs, and greater demand for critical care resources.

These findings highlight the need to strengthen the early management of obstetric emergencies, optimize perioperative care, and enhance timely stabilization to reduce the severity of illness at ICU admission and subsequently shorten the ICU stay.

### Factors associated with the clinical outcomes of maternal ICU admission

#### Factors associated with Mortality

Mechanical ventilation (p = 0.04) and inotropes and/or vasopressor use (p = 0.022) were significantly associated with mortality in this study. This finding is consistent with reports from studies done in Malawi by Adeniran et al (47), India by Soni at al (48) and by Kumar et al (49) which both indicated that, mechanical ventilation was significantly associated with increased maternal mortality (p= 0.028), (p = <0.0001) and (p= 0.010) respectively. Similarly, in this study, the use of inotropes and/or vasopressors was significantly associated with mortality (p = 0.022). This finding is also consistent with the findings of studies from India by Adeniran et al. (47) and Kumar et al. (49), which indicated an association between inotropes and/or vasopressor use and increased mortality (p = <0.001) and (p = <0.001), respectively. The observed significant association between the need for mechanical ventilation and the use of inotropes and/or vasopressors with maternal mortality indicates that these interventions are strong markers of critical illness and poor prognosis among women admitted to the ICU following obstetric surgery. Clinically, this underscores the importance of early recognition and timely management of deteriorating patients before progression to respiratory failure or hemodynamic instability, requiring advanced organ support.

#### Factors associated with prolonged ICU stay

Mechanical ventilation and blood and/or blood product transfusion were significantly associated with prolonged ICU stay in both univariate and multivariable modified Poisson regression analyses. After adjusting for potential confounders, mothers who required mechanical ventilation had a significantly higher prevalence of prolonged ICU stays than those who did not require ventilatory support (p = 0.025). This finding aligns with the study conducted in China by Zhao et al. (50), who reported that mechanical ventilation was strongly associated with prolonged ICU stay in postpartum patients. Similarly, patients who received blood and/or blood products had a significantly higher prevalence of prolonged ICU stay than those who did not receive blood or blood products (p = 0.003). This finding aligns with the study conducted in Turkey by Agacayak et al. (33), who reported that blood product transfusion was the most effective independent parameter affecting the duration of stay in the ICU (p=0,001). The association of mechanical ventilation and blood and/or blood product transfusion with prolonged ICU stay suggests that these interventions are markers of severe illness and increased physiological instability among obstetric patients. Clinically, this highlights the importance of early recognition and prompt management of complications, such as PPH and critical deterioration, to prevent progression to conditions requiring advanced support and reduce the duration of ICU stay.

#### Study limitations

i. The study was conducted at one tertiary hospital, which may limit generalizability to lower-level health facilities
ii. Another limitation is since the study is cross-sectional, makes it impossible to definitively determine causal-effect relationships between the variables studied
iii. The study did not assess long term outcomes to the participants

## CONCLUSION

The findings of this study revealed that PPH was the most common indication for maternal ICU admission following obstetric surgery, accounting for 46.1% of admissions. Furthermore, Prolonged ICU stay was observed in 40.9% of patients, while the overall maternal mortality was 16.5%. Mechanical ventilation and blood and/or blood product transfusion were significantly associated with prolonged ICU stay. In addition, mechanical ventilation and the use of inotropes and/or vasopressors were significantly associated with maternal mortality. Overall, these findings indicate a substantial burden of severe maternal morbidity and mortality among women requiring ICU admission after obstetric surgeries. Clinically, the predominance of hemorrhagic complications and the association of advanced life support interventions with poor outcomes highlight the need for early recognition of risk factors of PPH during ANC visits, timely recognition and management of obstetric emergencies, optimized perioperative monitoring, and strengthened critical care services to improve maternal outcomes.

## Data Availability

The data underlying the findings of this study are derived from routinely collected clinical information from medical records, ICU registers, anaesthesia notes, surgical notes, and ICU management charts of obstetric ICU patients following obstetric surgery. Public deposition of the complete participant-level dataset is therefore restricted by the ethical approval and institutional data-governance requirements governing this study. The study was approved by the Muhimbili University of Health and Allied Sciences Research Ethics Committee (MUHAS-REC-04-2025-2788). De-identified data sufficient to reproduce the analyses reported in this article may be made available to qualified researchers following review and approval by the relevant institutional authorities and ethics committee, and subject to applicable data-sharing and data-transfer requirements. Requests for access should be directed to the Muhimbili University of Health and Allied Sciences Research Ethics Committee or the appropriate institutional data-access authority at MUHAS. Data access is not controlled solely by the study authors.

